# Immune checkpoint inhibitors combined with antiangiogenic drugs in the treatment of unresectable locally advanced / distant metastatic esophageal squamous cell carcinoma: a systematic review protocol

**DOI:** 10.1101/2023.05.20.23286898

**Authors:** Juanjuan Lu, Honglin Li, Sisi Chang, YahuiZhu, Fuyan Gao, Shuai Shao, Chunzheng Ma

**Author notes:** Corresponding author: Honglin Li, female, Doctor degree, attending physician, master supervisor, Department of Oncology, Henan Hospital of Traditional Chinese Medicine,. Author: Juanjuan Lu, female, postgraduate student, Henan University of Traditional Chinese Medicine (Second Clinical Medical College),.

## Abstract

**Background and purpose:** Esophageal cancer has the seventh highest incidence and sixth highest fatality rate of all malignant tumors. It’s a type of cancer that has a greater fatality rate than morbidity. In the treatment of advanced esophageal and squamous cell carcinoma, targeted and immune checkpoint inhibitors have showed promising outcomes in recent years. The aim of this research was to examine the effectiveness and safety of immune checkpoint inhibitors in combination with antiangiogenic medicines in the treatment of unresectable patients with locally progressed / distant metastatic esophageal squamous cell carcinoma.

**Methods:** Before April 15, 2022, randomized controlled trials of immune checkpoint inhibitors combined with anti-angiogenic drugs in the treatment of esophageal cancer will be searched on PubMed, Cochrane Library, EMBASE, WebofScience, China Knowledge Network, Wanfang Data Knowledge Service Platform, and VIP. The two authors independently extracted the data, checked the data, and used the “bias risk” tool in the Cochrane intervention system evaluation manual to independently evaluate the bias included in the study. When the extracted data were similar enough, the Revman5.4 software was used for meta analysis. If the summary data could not be collected for meta analysis, the results would be summarized in a narrative way.

**Discussion:** This paper introduces in detail the systematic review and design method of immune checkpoint inhibitors mixture with antiangiogenic drugs in the treatment of unresectable locally advanced / distant metastatic esophageal squamous cell carcinoma.

**Systematic review registration:** The review protocol has been registered in PROSPERO CRD42022324666

## Background

Esophageal cancer (EC) is a type of malignant tumor principally involving the squamous epithelium and columnar epithelium of the esophagus, EC has the seventh incidence rate and the sixth mortality rate^[1]^. The prime pathologic patterns of EC are squamous cell carcinoma and adenocarcinoma, while esophageals squamous cell carcinoma (ESCC)make up more than 90% of all EC ^[2-3]^. According to statistics, there were only 572000 new cases of EC from poles to poles in 2018, of which ESCC accounted for 482000 cases ^[4]^. In recent years, the overall morbidity of EC has decreased, but the mortality rate remains high. The total five-year survival rate of patients with EC is only 16%, while the median survival time is less than 1 year ^[5-6]^.

The conventional treatment for EC is surgical resection or subresection combined with lymph node dissection, although more than half of ESCC patients are progressed at the time of treatment and cannot receive routine treatment [7-8]. Patients with EC have a 5-year survival rate of fewer than 20% when treated with radiation, chemotherapy, or surgical combination treatment [9]. This demonstrates that there is a critical need to investigate new treatment options for unresectable locally progressed / distant metastatic ESCC. In the treatment of advanced ESCC, targeted and immune checkpoint inhibitors (ICI)have showed promising outcomes in recent years [10].Wang,F et al. ^[11]^ showed that canrelizumab combined with apatinib as a second-line treatment of ESCC achieved a good clinical efficacy, and this is also the first randomized controlled trial of the programmed cell death-1 receptor (PD-1) inhibitors combined with anti-angiogenesis inhibitors in the therapy of terminal ESCC.

However, there is yet little evidence to support the use of ICI in combination with antiangiogenic medicines in the treatment of advanced esophageal squamous cell carcinoma. In this study, we’ll look at progression-free survival (PFS), overall survival (OS), objective remission rate (ORR), quality of life, and adverse events, as well as the efficacy and safety of ICI paired with anti-angiogenic medicines. To provide clinicians with a solid foundation.

## Method/design

The method we used will be consistent with the PRISMA-P Statement [12] (see additional file 1) report, the preferred reporting item for the Systems evaluation and Meta-analysis program, and has been registered with the International Prospective Systems Review Registry (PROSPERO), registration number CRD42022324666.

We will use the PICO principle to describe the research problem, as shown in Table 1.

**Table 1:**

|  |  |
| --- | --- |
| P | Patients with unresectable locally advanced/distant metastatic esophageal squamous cell carcinoma confirmed by histology or cytology |
| I | Immune checkpoint inhibitor in combination with antiangiogenic drugs |
| C | chemotherapy |
| O | Overall survival, progression-free survival |
| Immune checkpoint inhibitors include: PD-1 inhibitors: nivolumab, pembrolizumab, toripalizumab, sintilimab, camrelizumab, tislelizumab Anti-, Pienpilimumab; PD-L1 Inhibitors: Durvalumab, Atezolizumab, Avaluzumab; CTLA-4 Inhibitor: Ipilimumab. Anti-angiogenic drugs include: bevacizumab, recombinant human endostatin, sorafenib, sunitinib, apatinib, anlotinib, fruquintinib, ramucirumab. Chemotherapy drugs include, but are not limited to: fluorouracil, cisplatin, taxane, docetaxel, oxaliplatin, leucovorin, and epirubicin, alone or in combination. |  |

### The study’s inclusion as well as exclusion criteria

The following criteria will be met by the researches included in this evaluation:

1. Patients with unresectable advanced or metastatic ESCC diagnosed histologically or cytologically;
2. Patients were treated with ICI and antiangiogenic agents;
3. At least one of the following outcomes should be reported: OS, PFS, ORR, quality of life assessment, and adverse reactions.
4. Randomized controlled trials.

The following criteria will be excluded:

1. duplicate publication of data;
2. Outcome indicators are not clear or cannot be combined;
3. Small cell esophageal carcinoma, esophageal adenocarcinoma, or other mixed carcinomas proved by histology or cytology.
4. Animal tests.

### Outcome measures of the study

The ultimate goal of cancer treatment is to help patients manage the progression of the disease and thus live longer. So we chose two primary endpoints:

1. PFS: the time from randomization to disease progression or death from any cause.
2. OS: time from randomization to death.

Secondary outcome measures included:

1. ORR: the proportion of patients whose tumor shrank by a certain amount for a certain period of time.
2. Quality of life evaluation: Karnofsky score was adopted.
3. Adverse reactions.

### Retrieval strategy

We will search relevant studies published before April 15, 2022 in PubMed, Cochrane Library, EMBASE, Web of Science, CNKI, Wan fang Data Knowledge Service Platform, and VIP through the combination of subject words and free words, without language restrictions. Also, we will re-conduct the search before the final analysis. Additional file 2 for more detailed Pubmed database search strategy.

### Study selection

The two authors (HLL,JJL) will independently search the literature, select the potential studies that meet the inclusion criteria, import the literature management software NoteExpress to delete the duplicate literatures, then read the literature title and abstract, and include the preliminary qualified literatures according to the established criteria. Then, after reading the full text, the literatures will be screened again and the reasons for the exclusion of the full text will be recorded. Then cross-check. If the two parties disagree, we will communicate with the third author (SSC) until consensus is reached to ensure that the included literature is comprehensive and accurate.We will document the selection process in detail in the PRISMA flowchart, see additional file 3.

### Data extraction and management

We will extract the data eventually included in the study according to the pre-designed inclusion study feature table (Table 2). Two authors (HLL,JJL) will extract the data independently and record the following specific data in the table.

1. Study characteristics and methods: first author, publication date, randomization, allocation hiding, and median follow-up time.
2. Participants: Total number of participants in each group, country, gender, age, tumor stage, pathological diagnosis, PD-L1 protein expression level.
3. Intervention: specific treatment plan.
4. Other data: OS, PFS, ORR, adverse reactions, quality of life assessment, 95% confidence interval.

**Table 2:**
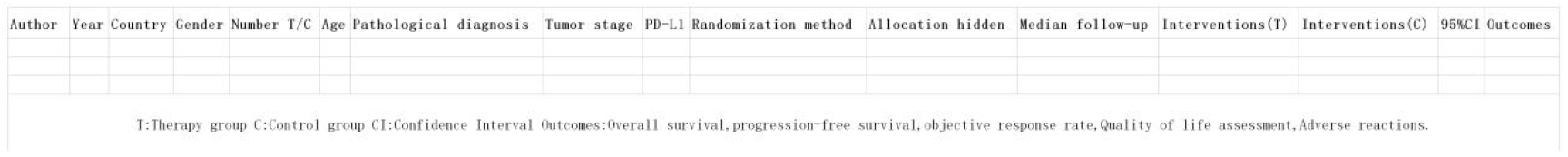

If the data in the report is missing or unclear, the first author will be contacted by email for specific data.

### Quality evaluation method

Two authors (HLL,JJL) independently assessed the bias of the included studies using the “risk of bias” tool in the Handbook of Systematic Review of Cochrane Interventions ^[13]^.A third author (SSC) will be consulted when differences arise.

Each of the following areas will be evaluated: random sequence generation (selection bias);Allocation hiding (selection bias);Blindness of subjects and intervention implementors (implementation bias);Blind method of outcome evaluators (detection bias);Results data were incomplete (lost to follow-up bias);Selective reporting (reporting bias);Other biases (common other biases: availability of clear inclusion/exclusion criteria; Whether to calculate the sample size; Sample size balance between groups; Whether baseline data are comparable; Financial support and conflict of interest).Each area will be assessed for high risk, low risk, or uncertain risk bias. The evaluation results will be reported on the Risk quality assessment chart.

### Statistical analysis

We will use Revman5.4 software to analyze the data. We will conduct meta analysis only if the participants, interventions and outcome measurements included in the study are similar enough. For the analysis of event occurrence time results, we use the Hazard ratio (HR) and its 95% confidence interval (CI)as the effect measure of each study ^[14]^. For binary data, we will calculate relative risk (RR) and 95% CI based on the analysis of the number of events and the number of participants in the intervention group and the control group. For consecutive results, if all the results are measured on the same scale, we will summarize the mean difference (MD) between the treatment groups. We will use Cochran Q and *I*^*2*^ statistics to assess the statistical heterogeneity between trials. When *P* < 0.10 or *I*^*2*^ > 50%, it indicates that there is obvious heterogeneity ^[15]^, then choose the random effect model to analyze the data, otherwise we will use the fixed effect model. If the same study results use different measurement methods, we will use the inverse variance method to calculate standardized mean difference(SMD) and 95%CI, and we will analyze the data based on the mean, standard deviation and number of participants between the intervention group and the control group to calculate the mean difference and 95%CI between the treatment group. If we are unable to count the aggregate data for meta analysis, we will conduct a narrative summary of the results. When there is significant heterogeneity, we will do a subgroup analysis of the main outcome indicators, such as the selection of immune checkpoint inhibitors. When the sample size is sufficient to support the analysis (randomized controlled trial > 10), we will investigate the publication bias of the funnel chart and do sensitivity analysis.

### Evidence quality assessment

The included outcome indicators were evaluated according to the GRADE tool. The GRADE system evaluates evidence grades based on five grading factors (bias risk, indirectness, imprecision, inconsistency, and publication bias), with drop 1 grade to medium quality, drop 2 grade to low quality, drop 3 grade to very low quality. High quality evidence is strong recommendation, low quality and very low quality is weak recommendation. Finally, the quality of evidence is divided into four levels: high quality, medium quality, low quality and very low quality. High quality evidence is strong recommendation, low quality and very low quality is weak recommendation.

## Discussion

EC is a global medical and health problem. It is an aggressive malignant tumor. Surgical resection is a radical treatment. However, most of the patients have local advanced/distant metastasis and cannot be treated by surgery when they come to the clinic. Therefore, the prognosis is poor and new treatment methods are urgently needed. Targeted and ICI have been shown to be efficacious in the management of advanced esophageal squamous cell carcinoma in recent years. Wang, F et al. [11] were the first to study the combination of PD-1 inhibitors and anti-angiogenic inhibitors in the treatment of advanced ESCC, and the clinical results were encouraging, which would be the beginning of a new therapy. Currently, ICI combined with antiangiogenic agents in the therapy of terminal esophageal squamous cell carcinoma are limited, and there is no systematic evaluation of this new therapy. This study will be the first systematic review of immunocheckpoint inhibitors combined with antiangiogenic agents in the treatment of unresectable locally advanced/distant metastatic esophageal squamous cell carcinoma.

This article follows PRISMA-P standards and has been registered with the International Registry for Prospective Systems Review (PROSPERO), registration number CRD42022324666. The research and design methods of system evaluation and meta-analysis are introduced in detail, including inclusion/exclusion criteria, outcome indicators, search strategies, study selection, data extraction and management, quality assessment methods, statistical analysis, and the GRADE system evaluation will be used to evaluate the grade of outcome indicators. Our primary objective is to provide reliable data on the efficacy and safety of immunocheckpoint inhibitors combined with antiangiogenic agents in the treatment of advanced esophageal squamous cell carcinoma for scientific use in clinical studies.

## Data Availability

All data produced in the present work are contained in the manuscript

## Abbreviations

EC: Esophageal cancer
ESCC: Esophageals squamous cell carcinoma
ICI: Immune checkpoint inhibitors
PD-1: Programmed cell death-1 receptor
PFS: Progression-free survival
OS: Overall survival
ORR: Objective remission rate
PRISMA-P: Preferred Reporting Items for Systematic Review and Meta-Analysis Protocols
CNKI: China National Knowledge Internet
HR: Hazard ratio
CI: Confidence interval
RR: Relative risk
MD: Mean difference
SMD: Standardized mean difference

## Declarations

### Ethics approval and consent to participate

Not applicable.

### Consent for publication

Not applicable.

### Availability of data and materials

Not applicable.

### Authors’ contributions

This study was primarily conceived by HLL, JJL, and SSC, with JJL writing the first draft of the protocol manuscript and rigorously revising it by HLL. SSC, YHZ, FYG, SS registered the protocol in the PROSPERO database. CZM reviewed the manuscript. All authors have read and approved this agreement for publication.

## Acknowledgements

Not applicable.

## Authors’ information

Corresponding author:HLL

