## Additional file 2 for "Immune checkpoint inhibitors combined with antiangiogenic drugs in the treatment of unresectable locally advanced / distant metastatic esophageal squamous cell carcinoma: a systematic review protocol"

| Database | Retrieval Strategy |
| --- | --- |
| PUBMED | Search:****("EsophagealNeoplasms"[Mesh])OR((((((((((((((((EsophagealNeoplasm[Title/Abstract]) OR (Neoplasm, Esophageal[Title/Abstract])) OR (Esophagus Neoplasm[Title/Abstract])) OR (Esophagus Neoplasms[Title/Abstract])) OR (Neoplasm, Esophagus[Title/Abstract])) OR (Neoplasms, Esophagus[Title/Abstract])) OR (Neoplasms, Esophageal[Title/Abstract])) OR (Cancer of Esophagus[Title/Abstract])) OR (Cancer of the Esophagus[Title/Abstract])) OR (Esophagus Cancer[Title/Abstract])) OR (Cancer, Esophagus[Title/Abstract])) OR (Cancers, Esophagus[Title/Abstract])) OR (Esophagus Cancers[Title/Abstract])) OR (Esophageal Cancer[Title/Abstract])) OR (Cancer, Esophageal[Title/Abstract])) OR (Cancers, Esophageal[Title/Abstract])) AND**** Search:****("Immune Checkpoint Inhibitors"[Mesh]) OR (((((((((((((((((((((((((((((((((Checkpoint Inhibitors, Immune[Title/Abstract]) OR (Immune Checkpoint Inhibitor[Title/Abstract])) OR (Checkpoint Inhibitor, Immune[Title/Abstract])) OR (Immune Checkpoint Blockers[Title/Abstract])) OR (Checkpoint Blockers, Immune[Title/Abstract])) OR (Immune Checkpoint Blockade[Title/Abstract])) OR (Checkpoint Blockade, Immune[Title/Abstract])) OR (Immune Checkpoint Inhibition[Title/Abstract])) OR (Checkpoint Inhibition, Immune[Title/Abstract])) OR (PD-L1 Inhibitors[Title/Abstract])) OR (PD L1 Inhibitors[Title/Abstract])) OR (PD-L1 Inhibitor[Title/Abstract])) OR (PD L1 Inhibitor[Title/Abstract])) OR (Programmed Death-Ligand 1 Inhibitors[Title/Abstract])) OR (Programmed Death Ligand 1 Inhibitors[Title/Abstract])) OR (PD-1-PD-L1 Blockade[Title/Abstract])) OR (Blockade, PD-1-PD-L1[Title/Abstract])) OR (PD 1 PD L1 Blockade[Title/Abstract])) OR (CTLA-4 Inhibitors[Title/Abstract])) OR (CTLA 4 Inhibitors[Title/Abstract])) OR (CTLA-4 Inhibitor[Title/Abstract])) OR (CTLA 4 Inhibitor[Title/Abstract])) OR (Cytotoxic T-Lymphocyte-Associated Protein 4 Inhibitors[Title/Abstract])) OR (Cytotoxic T Lymphocyte Associated Protein 4 Inhibitors[Title/Abstract])) OR (Cytotoxic T-Lymphocyte-Associated Protein 4 Inhibitor[Title/Abstract])) OR (Cytotoxic T Lymphocyte Associated Protein 4 Inhibitor[Title/Abstract])) OR (PD-1 Inhibitors[Title/Abstract])) OR (PD 1 Inhibitors[Title/Abstract])) OR (PD-1 Inhibitor[Title/Abstract])) OR (Inhibitor, PD-1[Title/Abstract])) OR (PD 1 Inhibitor[Title/Abstract])) OR (Programmed Cell Death Protein 1 Inhibitor[Title/Abstract])) OR (Programmed Cell Death Protein 1 Inhibitors[Title/Abstract]))AND****Search: ****((((((((((((((((((((((((((((((((((((((((((((((((((((((((((Angiogenesis Inhibitor[Title/Abstract]) OR (Inhibitor, Angiogenesis[Title/Abstract])) OR (Angiogenetic Antagonist[Title/Abstract])) OR (Antagonist, Angiogenetic[Title/Abstract])) OR (Angiogenetic Antagonists[Title/Abstract])) OR (Antagonists, Angiogenetic[Title/Abstract])) OR (Angiogenetic Inhibitor[Title/Abstract])) OR (Inhibitor, Angiogenetic[Title/Abstract])) OR (Angiogenetic Inhibitors[Title/Abstract])) OR (Angiogenic Antagonists[Title/Abstract])) OR (Angiogenic Antagonist[Title/Abstract])) OR (Antagonist, Angiogenic[Title/Abstract])) OR (Angiogenic Inhibitor[Title/Abstract])) OR (Inhibitor, Angiogenic[Title/Abstract])) OR (Angiostatic Agent[Title/Abstract])) OR (Agent, Angiostatic[Title/Abstract])) OR (Anti-Angiogenetic Agent[Title/Abstract])) OR (Agent, Anti-Angiogenetic[Title/Abstract])) OR (Anti Angiogenetic Agent[Title/Abstract])) OR (Angiogenic Inhibitors[Title/Abstract])) OR (Angiostatic Agents[Title/Abstract])) OR (Agents, Angiostatic[Title/Abstract])) OR (Antagonists, Angiogenic[Title/Abstract])) OR (Anti-Angiogenetic Agents[Title/Abstract])) OR (Agents, Anti-Angiogenetic[Title/Abstract])) OR (Anti Angiogenetic Agents[Title/Abstract])) OR (Anti-Angiogenic Drugs[Title/Abstract])) OR (Anti Angiogenic Drugs[Title/Abstract])) OR (Drugs, Anti-Angiogenic[Title/Abstract])) OR (Antiangiogenic Agents[Title/Abstract])) OR (Agents, Antiangiogenic[Title/Abstract])) OR (Inhibitors, Angiogenesis[Title/Abstract])) OR (Inhibitors, Angiogenetic[Title/Abstract])) OR (Inhibitors, Angiogenic[Title/Abstract])) OR (Inhibitors, Neovascularization[Title/Abstract])) OR (Neovascularization Inhibitors[Title/Abstract])) OR (Anti-Angiogenic Drug[Title/Abstract])) OR (Anti Angiogenic Drug[Title/Abstract])) OR (Drug, Anti-Angiogenic[Title/Abstract])) OR (Neovascularization Inhibitor[Title/Abstract])) OR (Inhibitor, Neovascularization[Title/Abstract])) OR (Antiangiogenic Agent[Title/Abstract])) OR (Agent, Antiangiogenic[Title/Abstract])) OR (Angiogenesis Factor Inhibitors[Title/Abstract])) OR (Factor Inhibitors, Angiogenesis[Title/Abstract])) OR (Inhibitors, Angiogenesis Factor[Title/Abstract])) OR (Angiogenesis Factor Inhibitor[Title/Abstract])) OR (Factor Inhibitor, Angiogenesis[Title/Abstract])) OR (Inhibitor, Angiogenesis Factor[Title/Abstract])) OR (Anti-Angiogenesis Effect[Title/Abstract])) OR (Anti Angiogenesis Effect[Title/Abstract])) OR (Effect, Anti-Angiogenesis[Title/Abstract])) OR (Antiangiogenesis Effect[Title/Abstract])) OR (Effect, Antiangiogenesis[Title/Abstract])) OR (Antiangiogenesis Effects[Title/Abstract])) OR (Effects, Antiangiogenesis[Title/Abstract])) OR (Anti-Angiogenesis Effects[Title/Abstract])) OR (Anti Angiogenesis Effects[Title/Abstract])) OR (Effects, Anti-Angiogenesis[Title/Abstract])**** |
