## Additional file 3 for "Immune checkpoint inhibitors combined with antiangiogenic drugs in the treatment of unresectable locally advanced / distant metastatic esophageal squamous cell carcinoma: a systematic review protocol"

**PRISMA 流程图**

Records identified through Database searching (n=)

Additional records identified through other sources (n=)

Duplicate records removed(n=)

Preliminary included literature (n=)

Records with full-test reviewed roughly (n=)

Records included in review

(n=)

Reports excluded for reported studies or studies with poor quality（n=）

Reports excluded for no full tests or not related obviously（n=）
